# Causal evidence for an ApoB-independent metabolomic risk profile associated with coronary artery disease

**DOI:** 10.1101/2023.03.01.23286619

**Authors:** Linjun Ao, Diana van Heemst, J. Wouter Jukema, Patrick C.N. Rensen, Ko Willems van Dijk, Raymond Noordam

## Abstract

**Background and aims:** 1-H nuclear magnetic resonance (^1^H-NMR) metabolomic measures in plasma have yielded significant insight into the pathophysiology of cardiometabolic disease, but their interrelated nature complicates causal inference and clinical interpretation. This study aimed to investigate the associations of unrelated ^1^H-NMR metabolomic profiles with coronary artery disease (CAD), type 2 diabetes (T2D) and ischemic stroke (ISTR).

**Methods:** Principal component (PC) analysis was performed on 168 ^1^H-NMR metabolomic measures in 56,712 unrelated European participants from UK Biobank to retrieve unrelated PCs, which were used in multivariable-adjusted cox-proportional hazard models and genome-wide association analyses for Mendelian Randomization (MR). Two-sample MR analyses were conducted in three non-overlapping databases which were subsequently meta-analysed, resulting in combined sample sizes of 755,481 (128,728 cases), 1,017,097 (121,977 cases), and 1,002,264 (56,067 cases) for CAD, T2D, and ISTR, respectively.

**Results:** We identified six PCs which collectively explained 88% of the total variance. For CAD in particular, results from both multivariable-adjusted and MR analyses were generally directionally consistent. The pooled odds ratios (ORs) [95% CI] of per one-SD increase in genetically-influenced PC1 and PC3 (both characterized by distinct ApoB-associated lipoprotein profiles) were 1.04 [1.03, 1.05] and 0.94 [0.93, 0.96], respectively. In addition, the pooled OR for CAD of PC4, characterized by simultaneously decreased small HDL and increased large HDL, and independent of ApoB, was 1.05 [1.03, 1.07]. For the other outcomes, PC5 (characterized by increased amino acids) was associated with a higher risk of T2D and ISTR.

**Conclusions:** This study highlights the existence of an ApoB-independent lipoprotein profile driving CAD. Interestingly, this profile is characterized by a distinctive HDL sub-particle distribution, providing evidence for a role of HDL in the development of CAD.

## Introduction

The effectiveness of lowering plasma low-density lipoprotein-cholesterol (LDL-C) to reduce coronary artery disease (CAD) risk is beyond doubt (1). Lowering of plasma triglyceride (TG) levels may also reduce CAD risk on top of LDL-C lowering (2, 3). These observational findings have been attributed to a reduction in apolipoprotein B (ApoB), which has been suggested as the primary marker for cardiovascular disease risk independent of lipid content (cholesterol or TG) and type of ApoB-containing lipoprotein (cholesterol-rich LDL or TG-rich VLDL) (4-7). Although an inverse association between high-density lipoprotein-cholesterol (HDL-C) and cardiovascular disease risk has long been established in prospective studies (8, 9), Mendelian randomization (MR) studies (10, 11) and clinical trials (12, 13) have so far failed to convincingly support a causal role for HDL-C in cardiovascular disease risk.

Importantly, lipoprotein metabolism is a highly dynamic system via which lipids and specific apolipoproteins are passively and actively exchanged between the different lipoprotein classes in the course of their transport and metabolism within the circulation. Considering lipoprotein classes such as LDL or HDL in isolation disregards the intricate interdependence of plasma lipoproteins. It would therefore be more appropriate to consider individual lipoprotein profiles as a whole, characterized by specific distributions of lipids and apolipoproteins over the different lipoprotein classes. Metabolomic platforms based on 1-H nuclear magnetic resonance (^1^H-NMR) imaging of plasma samples provide such individual profiles by generating detailed measures on the composition, size, number and distribution of the different lipoprotein classes in a sample (14). As an added bonus, other metabolomic measures, such as some amino acids are also reported. Analyses of metabolomic measures as intermediates between exposures and clinical outcomes, is a powerful approach to dissect complex etiologic mechanisms linking metabolic processes to disease (14-16).

The interrelated nature of the lipoproteins also makes it difficult to identify specific genetic instruments for a single lipid or lipoprotein species without pleiotropic effects on the other lipoprotein subclasses. When performing MR studies, this may lead to biased estimations of health effects (17). Here, we tested the hypothesis that individual ^1^H-NMR metabolomics profiles can be grouped into different overall patterns that are more or less independent from each other and that may have differential associations with cardiometabolic diseases. To address this hypothesis, we performed principal component analysis (PCA) on ^1^H-NMR metabolomics data from participants in the United Kingdom Biobank (UKB) who were free from disease at the time of sampling (18). The principal components (PCs) can be regarded as independent traits, characterized by a specific overall metabolomic profile, which were exploited to determine the associations with CAD, type 2 diabetes mellitus (T2D), and ischemic stroke (ISTR) and to triangulate findings from observational research through multivariable-adjusted regression and large-scale multicohort MR analyses (19).

## Methods

### Project design

In the present study, we performed PCA on ^1^H-NMR metabolomic measures and conducted prospective multivariable-adjusted regression analyses to investigate the associations between selected PCs and examined cardiometabolic diseases in UKB participants. In line with the principles of triangulation, by using selected PCs from UKB participants as exposure, we further conducted genome-wide association studies (GWAS) and subsequent MR studies to assess potential causal associations of selected independent PCs with examined cardiometabolic diseases.

### Study population

Prospective multivariable-adjusted regression analyses and genome-wide association analyses were performed in the UKB, which recruited 502,628 participants aged 40-69 years across the entire United Kingdom between 2006 and 2010. The UKB cohort study was approved by the North-West Multicentre Research Ethics Committee (MREC), and the access for information to invite participants was approved by the Patient Information Advisory Group (PIAG) from England and Wales. All participants provided electronic written informed consent for the study. A detailed description of the UKB cohort has been presented elsewhere (18).

Plasma metabolic biomarkers were measured from 121,726 randomly selected UKB participants, 110,002 of whom had complete metabolomic measures and genetics data. To minimize ancestry and population stratification bias, we restricted the study population to 71,736 unrelated individuals of European ancestry, based on the estimated kinship coefficients for all pairs and the self-reported ancestral background (20). A subset of 57,846 participants free from CAD, T2D, ISTR and without taking cholesterol-lowering medication prior to the baseline survey were then selected for further studies. Finally, a total of 56,712 participants with complete data on covariables, including age, sex, the Townsend deprivation index, smoking status, alcohol consumption, body mass index (BMI), blood pressure lowering medication, and fasting time, were eligible for this study.

### Profiling of metabolomic measures

The measurement of metabolomic data took place between June 2019 and April 2020 using a high throughput and validated ^1^H-NMR-metabolomics platform (Nightingale Health, Helsinki, Finland). Technical details of the platform and epidemiological applications have been reviewed previously (14, 21).

The 168 direct metabolomic measures, comprising apolipoproteins (n = 2), lipoprotein particle sizes and concentration (n = 7), lipoprotein (sub)classes (n = 98), cholesterol (n = 15), triglycerides (n = 4), phospholipids and other lipids (n = 8), total lipids (n = 4), glycolysis related metabolites (n = 4), inflammation (n = 1), fluid balance (n = 2), fatty acids (n = 9), amino acids (n = 10), and ketone bodies (n = 4), were included in this study. A full list of the measured 168 metabolites and their concentration characteristics in our study population is presented in Table S1.

### Statistical analysis

#### Principal component analysis

PCA is a method used for dimension reduction by projecting each data point onto a new orthogonal coordinate system while capturing as much of the variation as possible (22), and can thus be used to identify uncorrelated patterns of a large number of interrelated risk factors, also known as PCs. All 168 metabolomic measures from 56,712 participants were first transformed to approximate a normal distribution by inverse rank-based normal transformation and standardized with standard deviation one and mean zero, and then subjected to PCA. The correlations between metabolomic measures and PCs could be represented by loadings (22). For each participant, each PC score was calculated by summing the standardized measures weighted by the corresponding eigenvector values (22).

#### Prospective analyses

The prospective multivariable-adjusted analyses were performed in the 56,712 UKB participants. Outcome diagnoses were coded according to the International Classification of Diseases edition 10 (ICD-10) and were based on the date of the first occurrence. CAD is defined as angina pectoris (I20), myocardial infarction (MI) (I21 and I22), and acute and chronic ischemic heart disease (IHD) (I24 and I25); ISTR is defined as cerebral infarction (I63); T2D is based on “non-insulin-dependent diabetes mellitus (E11)”. The sources of these variables are from hospital admissions (through linkage with the medical records from the National Health Service), primary care, death register, and through self-report. Based on the date of first appearance derived from the different information sources and the date of enrollment, we defined whether a case was prevalent (before enrollment) or incident (after enrollment). Outcomes in this prospective analysis were incident diseases during the time period from recruitment to January 1st, 2021. Follow-up time is computed from the baseline visit to the diagnosis of incident disease, loss-to-follow-up or death, or the end of the study period, whichever came first.

Three multivariable-adjusted Cox proportional hazard models were fitted to estimate hazard ratios (HRs) and corresponding 95% confidence intervals (95% CI) for the association between PCs and incident CAD, T2D, and ISTR: Model 1 was adjusted for age, sex, and the Townsend deprivation index; Model 2 was additionally adjusted for smoking status, alcohol consumption frequency, BMI, and blood pressure lowering medication; Model 3 was additionally adjusted for fasting time.

#### Mendelian randomization

MR uses genetic variants, typically single-nucleotide polymorphisms (SNPs), as instrumental variables (23, 24). MR studies depend on three main assumptions, notably: 1) the genetic variant must be associated with the exposure; 2) the genetic variant should not be associated with confounders; 3) the genetic variant affects the outcome only through the exposure. This study used the two-sample MR method, which requires that groups of participants in the gene-exposure association analysis and gene-outcome association analysis do not overlap (25-27).

##### Genotyping and genetic imputations

UK Biobank genotyping was conducted by Affymetrix using a bespoke BiLEVE Axium array for approximately 50,000 participants, and using the Affymetrix UK Biobank Axiom array for the remaining participants. All genetic data were quality controlled centrally by UK Biobank resources. More information on the genotyping processes can be found online (https://www.ukbiobank.ac.uk). Based on the genotyped SNPs, UK Biobank resources performed centralized imputations on the autosomal SNPs using the UK10K haplotype (28), 1000 Genomes Phase 3 (29), and Haplotype Reference Consortium reference panels (30). Autosomal SNPs were pre-phased using SHAPEIT3 and imputed using IMPUTE4. In total, ∼96 million SNPs were imputed.

##### Associations of genetic variants with exposure

GWAS were performed on each selected PC for the included 56,712 UKB individuals, using the software program GEM (version 1.4.2) (31), adjusted for age, sex, first 10 genetic PCs, and fasting time. SNPs with a minor allele frequency below 0.001 were removed. For each PC, genome-wide significant SNPs (*P* < 5*10^−8^) were selected, and then pruned to obtain independent instrumental variables by the TwoSampleMR and IeuGWASR packages, which use the PLINK clumping method with a clumping window of 10Mb and linkage disequilibrium r^2^<0.001 (32). To avoid potential residual pleiotropic effects, only SNPs without overlap among PCs were selected and subsequently used to extract the gene-outcome associations. The proportion of exposure variation explained by the genetic variants was depicted by the *R*^*2*^ statistic (33), and the potentially weak instrument bias was examined by the *F*-statistic, for which a threshold greater than 10 is conventionally considered sufficient for MR analysis (27).

##### Associations of genetic variants with outcome

Summary association statistics of the identified exposure-related SNPs with each outcome were estimated or extracted from 3 large databases, namely CARDIoGRAMplusC4D (34), DIAGRAM (DIAbetes Genetics Replication And Meta-analysis) consortium (35), and MEGASTROKE (36) for CAD, T2D, and ISTR, respectively, and UKB and FinnGen study for all three outcomes.

Participants from UKB were restricted to unrelated European-ancestry with the full released imputed genomics databases, and were not included in the variant-exposure association analysis. Outcomes in the MR analyses were prevalent or incident diseases, and details of all outcome diagnoses are described in the ‘prospective analysis’ subsection. In total, 34,299 cases and 227,723 controls for CAD, 22,659 cases and 239,363 controls for T2D, and 4,993 cases and 257,029 controls for ISTR were identified. Using the software program of GEM (version 1.4.2) (31), we performed the GWAS to assess the associations of genetic variants with T2D, CAD and ISTR, adjusting for age, sex, and the top 10 genetic principle components, and extracted the summary association statistics of identified exposure-related SNPs.

FinnGen is a public-private partnership research project launched in 2017, which covered the whole of Finland and combined genotype data from Finnish biobanks and digital health record data from Finnish health registries (37). In the FinnGen project and based on the ICD-10 (https://r7.risteys.finngen.fi/), major coronary heart disease (CHD), was defined as angina pectoris (I20), myocardial infarction (I21 to I23), ischemic heart diseases (I24 and I25), cardiac arrest (I46), and other cause unknown death or unattended death (R96 and R98); T2D was defined as non-insulin-dependent diabetes mellitus (E11); ISTR was defined as cerebral infarction (I63) and not specified as haemorrhage or infarction stroke (I64). Summary association statistics between identified exposure-related SNPs and the three outcomes were extracted from published FinnGen data freeze 7, which were based on 33,628 cases and 275,526 controls for major CHD, 44,313 cases and 255,449 controls for T2D, and 16,857 cases and 283,057 controls for ISTR.

The CARDIoGRAMplusC4D consortium assembled 60,801 cases and 123,504 controls from 48 studies for a GWAS meta-analysis of CAD, which was identified as an inclusive diagnosis of myocardial infarction, acute coronary syndrome, chronic stable angina, or coronary stenosis >50%. 77% of the participants were of European ancestry, 13% of South Asian ancestry, 6% of East Asian ancestry, and smaller samples were Hispanic and African Americans (34). The DIAGRAM consortium focuses on performing large-scale studies to characterise the genetic features of T2D. We selected the genome-wide association of T2D based on aggregated GWAS results from 31 studies for European-ancestry individuals with 55,005 cases and 400,308 controls, not including UKB samples (35). The large-scale MEGASTROKE consortium, launched by the International Stroke Genetics Consortium, releases the summary statistics from the 2018 meta-analysis of Genome-wide Association data in stroke and stroke subtypes with 34,217 ISTR cases and 406,111 controls (36).

We extracted summary association statistics of identified exposure-related SNPs with CAD, T2D, and ISTR from CARDIoGRAMplusC4D, DIAGRAM, and MEGASTROKE databases, respectively.

##### Associations of exposure with outcome

The associations of PCs with each study outcome from each database were estimated by the inverse-variance weighted (IVW) method, which combines the Wald ratio estimates (estimated association of genetic variants with outcome divided by estimated association of genetic variants with exposure) for individual genetic variant by a fixed-effect meta-analysis with inverse-variants weights (26, 27). Those estimates were expressed as log odds ratios (ORs) for the risk of CAD, T2D and ISTR for each PC per one-standard (one-SD) increase. We subsequently conducted meta-analyses for each PC to pool the estimates from the three outcome databases. The heterogeneity of the estimated ORs from three databases for each PC was represented by *I*^*2*^, and detected by the Cochran Q test (38).

Given that the IVW method assumes all genetic instruments are valid (e.g., no horizontal pleiotropy), we conducted sensitivity analyses using the weighted-median estimator and the MR-Egger method to assess whether IVW analyses were biased due to horizontal pleiotropy (39-41). Rather than taking a weighted mean of the ratio estimates as in the IVW method, the weighted-median estimator could still provide a consistent estimate of the causal effect even when up to 50% of the identified genetic variants are invalid IVs (39). In contrast to the IVW method, the MR-Egger method does not require a zero horizontal pleiotropy effect, and could detect pleiotropy by the intercept term (under the InSIDE assumption), which when different from zero indicates a bias in the IVW estimation (40, 41).

Except for the GWAS, all statistical analyses described above were performed in the R (version 4.0.2) software, with ‘prcomp’, ‘survival’ and ‘TwoSampleMR’ packages for PCA, Cox regression analyses and MR analyses, respectively.

## Results

### Principal component analysis

Metabolomic measures from 56,712 unrelated European-ancestry participants (57% women) with no history of CAD, T2D, stroke, and no cholesterol-lowering therapy at baseline were eligible for analyses in this study. PCA resulted in twelve PCs with eigenvalues above 1, which explained 93.8% of the variance of the original metabolomic data (Table S2). The loadings of the last six PCs (PC7 to PC12, Figure S1) were small (most absolute values < 0.3). Considering the combination of the eigenvalues-greater-than-one rule (42), explained variance and loading interpretability, the top six PCs with a cumulative explained variance of 87.95% were selected for further analyses (Figure 2).

**Figure 1.**
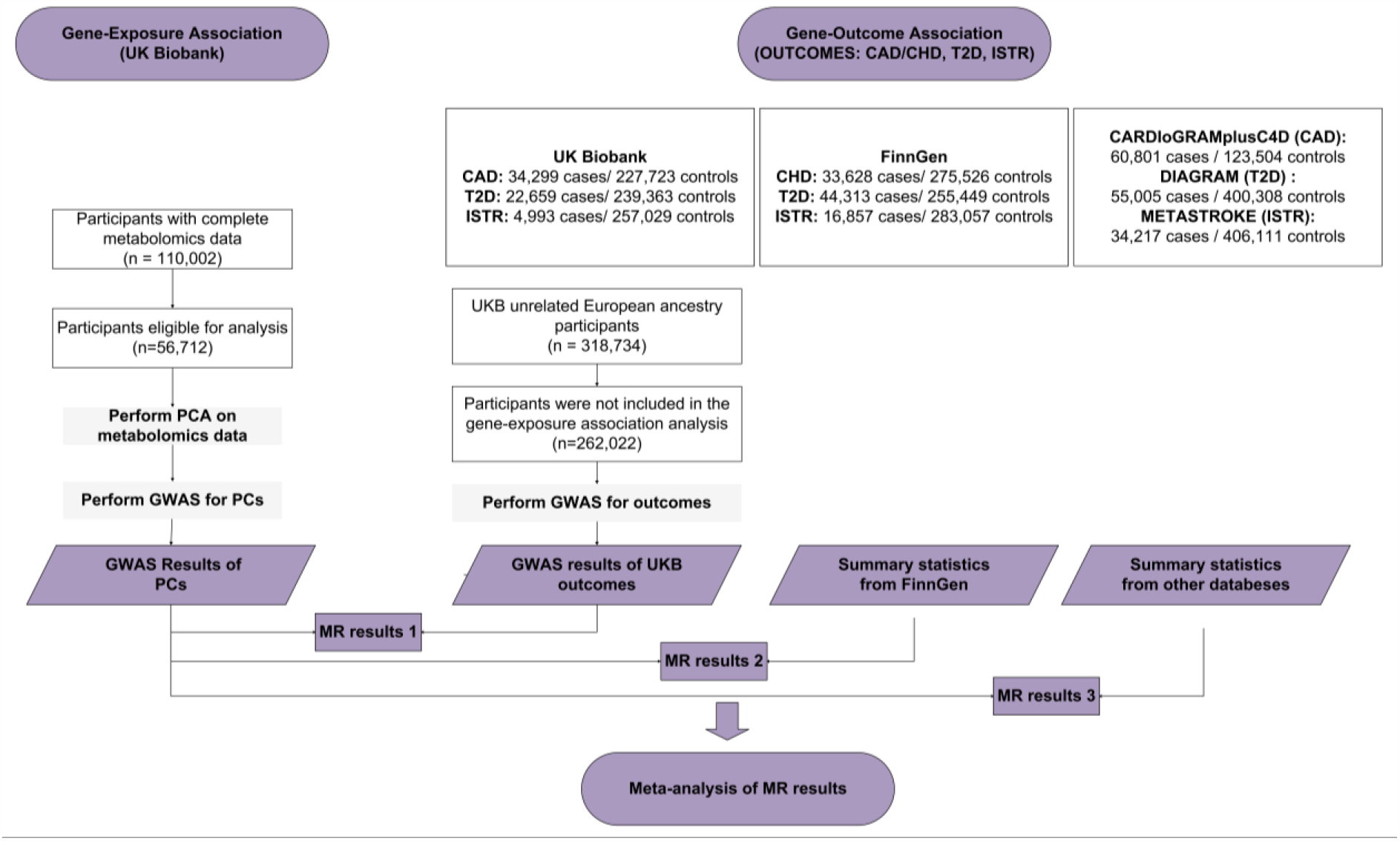
The overall workflow of Mendelian randomization analyses in the present study. CAD: coronary artery disease; CHD: coronary heart disease; T2D: type 2 diabetes; ISTR: ischemic stroke; PCA: principal component analysis; GWAS: genome-wide association study; MR: Mendelian Randomization.

**Figure 2.**
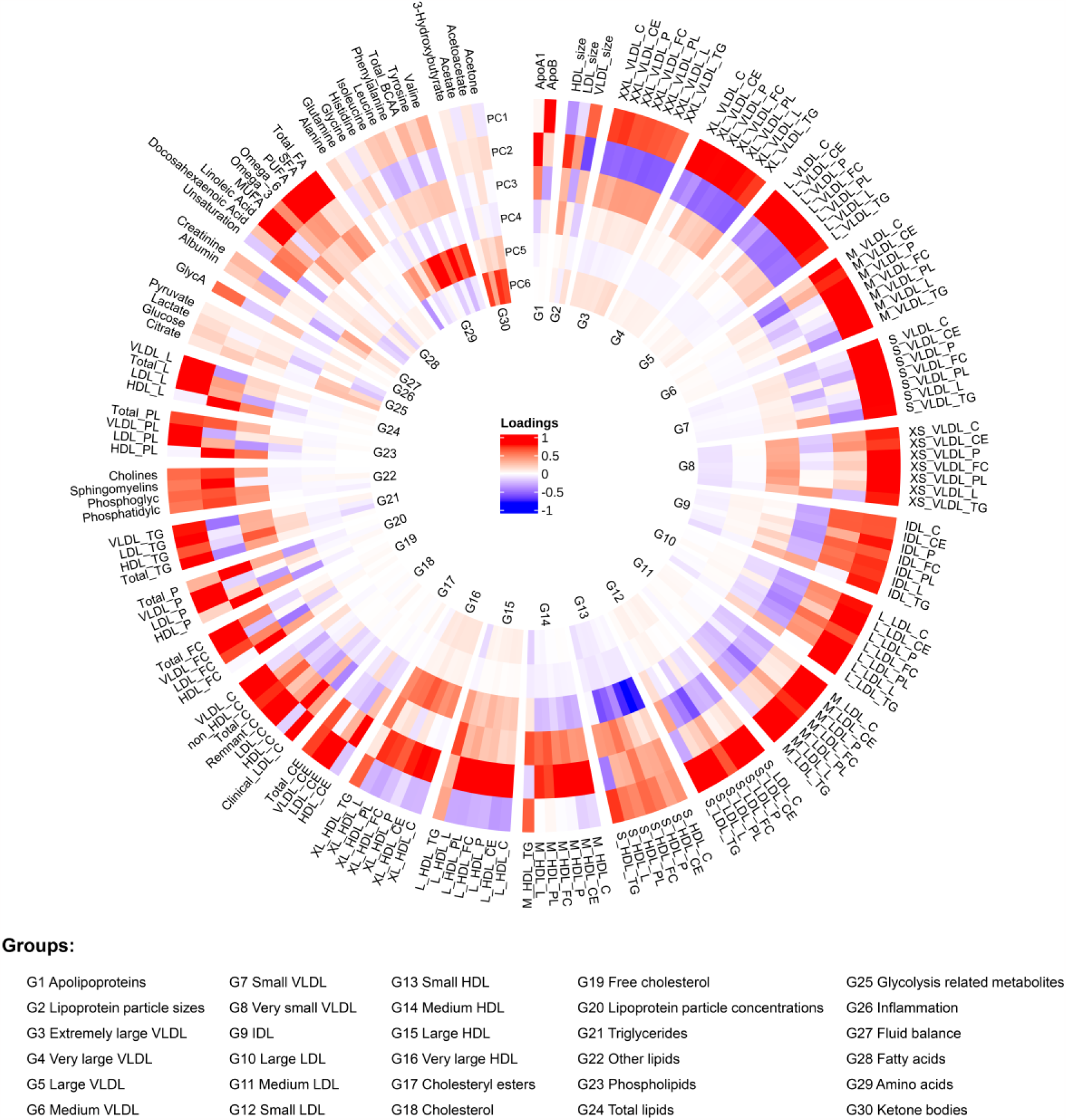
A circular loading plot of six selected principal components. From the outside to the inside, each circle represents the correlation between the respective principle components (PC) and 168 metabolomic measures. All metabolomic measures were divided into 30 groups and shown in clockwise order. Red or blue colour indicates the increase or decrease of metabolomic measures in PCs.

PC1 (46.7% variance) is mainly characterized by higher levels of ApoB, ApoB-containing lipoproteins and fatty acids. PC2 (22.5% variance) is mainly characterized by higher levels of apolipoprotein A1 (ApoA1), HDL particles and lower levels of VLDL particles. PC3 (9.4% variance) is characterized by lower levels of most ApoB-containing lipoproteins, but higher levels of HDL particles. PC4 (5.0% variance) is characterized by lower levels of small HDL particles and higher levels of very large HDL particles, independent of ApoB. PC5 (2.7% variance) is characterized by higher levels of amino acids, and PC6 (1.6% variance) is characterized by higher levels of ketone bodies.

### Prospective multivariable-adjusted regression analyses

The estimated multivariable-adjusted associations of each of the six PCs with the examined incident cardiometabolic diseases for UKB participants are presented in Figure 3. For the risk of CAD, the HRs [95% CI] for per one-SD increase in PC1, PC2, PC3 and PC4 were 1.02 [1.02, 1.03], 0.98 [0.97, 0.99], 0.98 [0.97, 0.99], and 1.02 [1.01, 1.03], respectively. For the risk of T2D, the HRs [95% CI] for per one-SD increase in PC1, PC2, PC3 and PC5 were 1.03 [1.02, 1.03], 0.93 [0.92, 0.94], 1.05 [1.04, 1.06], and 1.05 [1.03, 1.08], respectively. For the risk of ISTR, the HRs [95% CI] for per one-SD increase in PC4 and PC6 were 1.04 [1.01, 1.07] and 1.08 [1.03, 1.34], respectively.

**Figure 3.**
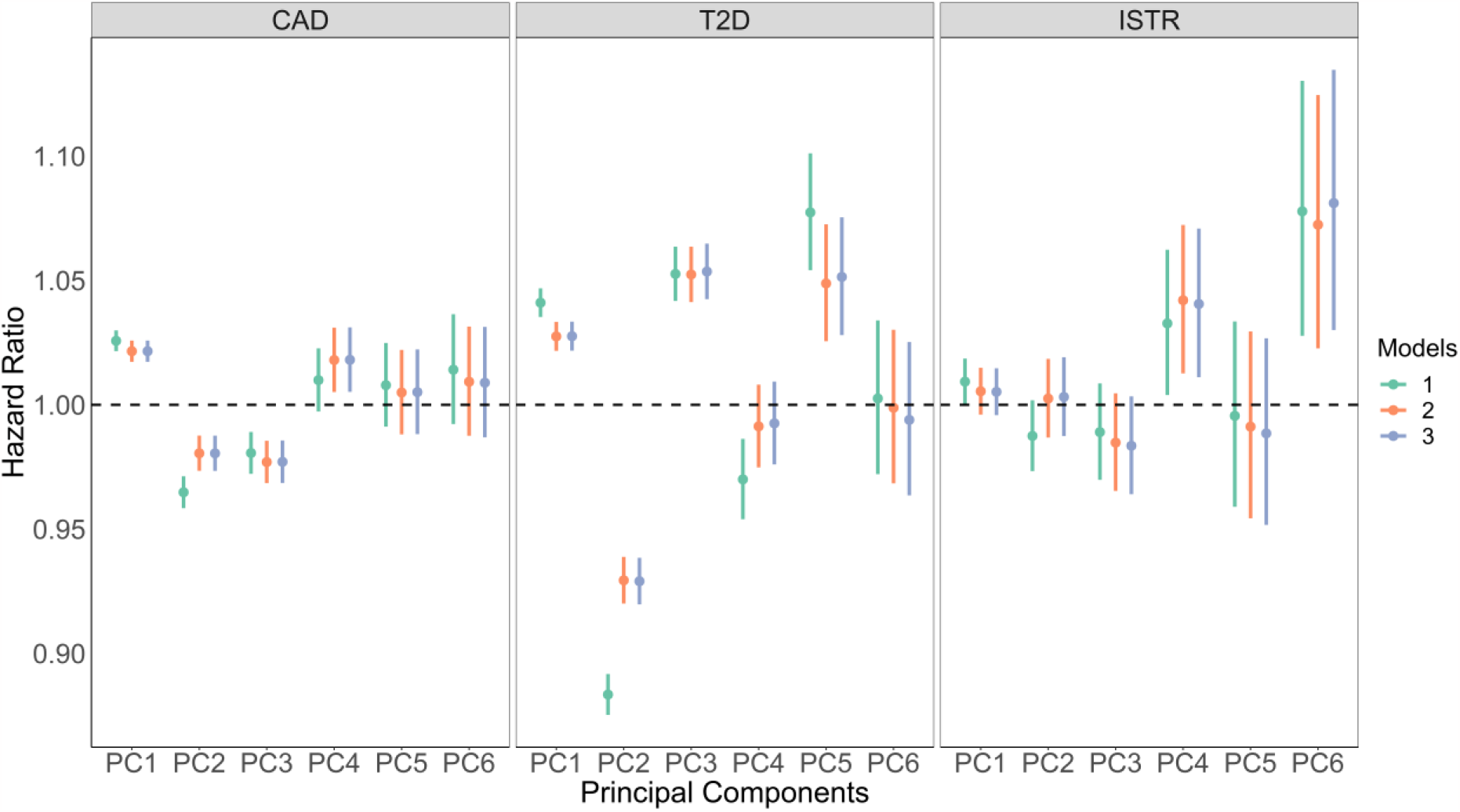
Hazard Ratios (95% CI) for incident coronary artery disease, ischemic stroke, and type 2 diabetes according to changes in principal components. CAD: coronary artery disease; ISTR: ischemic stroke; T2D: type 2 diabetes. Model 1 was adjusted for sex, age and Townsend index; Model 2 was model 1 additionally adjusted for smoking status, alcohol consumption frequency, BMI and blood pressure lowering medication; Model 3 was model 2 additionally adjusted for fasting time. The green, orange and blue lines indicate results based on models 1, 2 and 3, respectively.

### Mendelian randomization

A total of 150 independent SNPs were found to be significantly associated with the PCs, of which 41 SNPs, 37 SNPs, 31 SNPs, 22 SNPs, 11 SNPs, and 8 SNPs explained 7.3%, 9.0%, 7.5%, 9.0%, 0.9%, and 1.1% of the variation in PC1, PC2, PC3, PC4, PC5 and PC6, respectively. All *F* statistics were larger than 10. Supplementary tables provide the details of the independent and non-overlapping genetic variants, including their position, gene-exposure associations and corresponding *R*^*2*^ statistics and *F* statistics, and gene-outcome association (Table S3-8).

For the association of each PC with each outcome, Cochran Q statistics detected no heterogeneity (*P* values > 0.05) in the estimated ORs across the three outcome databases (Table S9). Figure 4 shows the estimated associations between each PC and each outcome from each database, and their pooled estimates across the three databases. For the risk of CAD, the pooled estimated ORs [95% CI] per one-SD increase in PC1, PC3 and PC4 were 1.04 [1.03, 1.05], 0.94 [0.93,0.96] and 1.05 [1.03, 1.07], respectively. For the risk of T2D, the pooled estimated ORs [95% CI] per one-SD increase in PC2 and PC5 were 0.98 [0.97,0.99] and 1.09 [1.02, 1.16], respectively. For the risk of ISTR, the pooled estimated ORs [95% CI] per one-SD increase in PC3 and PC5 were 0.97 [0.96,0.99] and 1.12 [1.07, 1.18], respectively.

**Figure 4.**
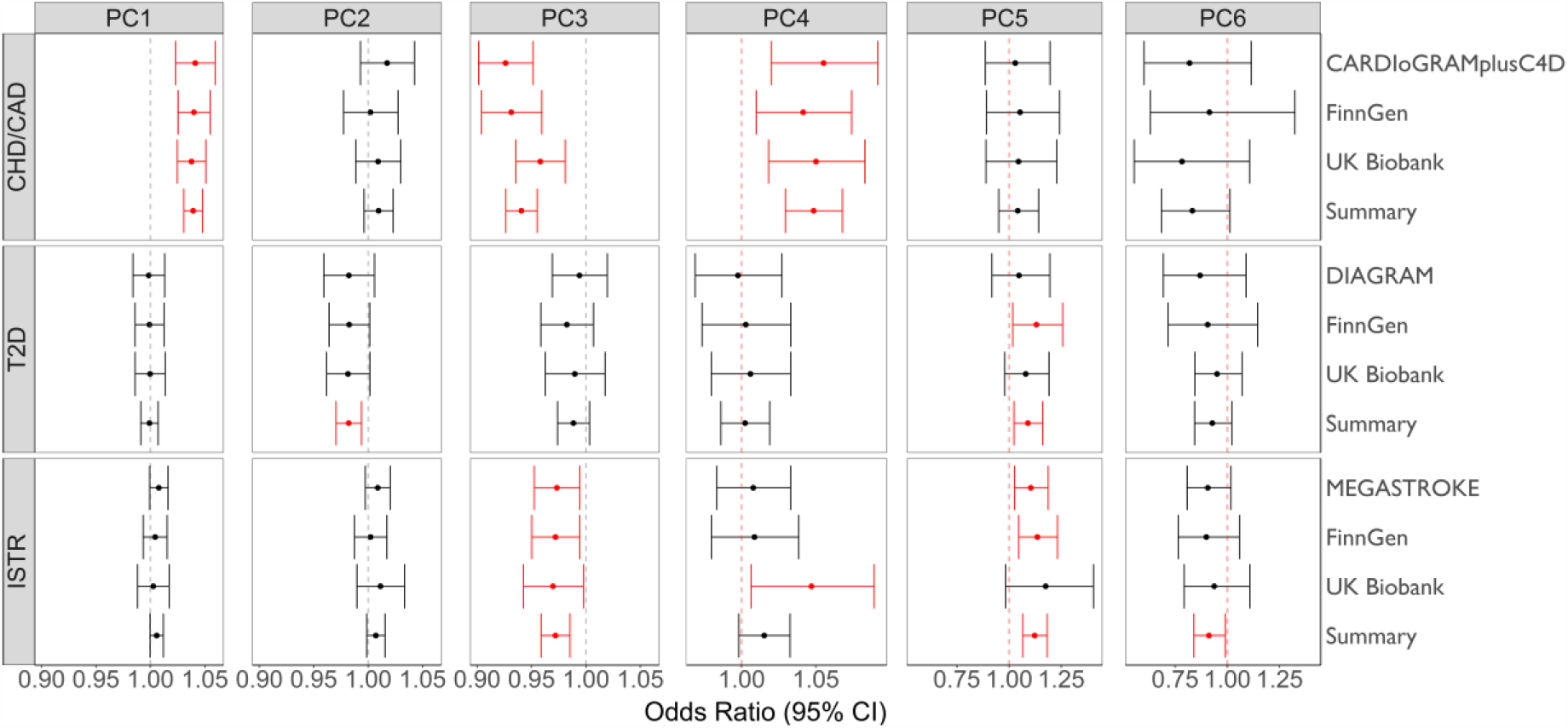
Odds ratios (95% CI) for the risk of cardiovascular disease, ischemic stroke, and type 2 diabetes estimated by inverse-variance weighted method for per one-SD increase in principal components. CAD: coronary artery disease; CHD: coronary heart disease; ISTR: ischemic stroke; T2D: type 2 diabetes. The right y-axis is labelled with the names of three data sources of the summary association statistics between genetic variants and outcome, and with ‘summary’ indicating the fixed-effect meta-analysis. Red lines (but not black lines) indicates estimated associations between principle components (PCs) and outcomes were statistically significant.

The estimated ORs based on the weighted-median estimator analyses were similar to those from IVW analyses (Table S9). No horizontal pleiotropic effect was detected according to the intercepts from MR-Egger, except for effects of PC5 on CHD from the FinnGen database (Table S10) and of PC1 and PC2 on T2D across three databases.

The loadings (Figure 2) and associations of PC1 and PC3 with CAD are in line with previous observations of ApoB as a major driver of CAD risk. Surprisingly, although the contribution of ApoB to PC4 is negligible, PC4 clearly associates with CAD. To assess the contribution of ApoB to CAD in PC4, we reperformed the MR analyses for the residual PC4, which was derived by regressing PC4 on ApoB. The MR result (Figure S2) using the IVW method showed that for the risk of CAD, the estimated pooled OR [95% CI] was 1.07 [1.03,1.11] for per one-SD increase in residual-PC4. This indicates that PC4 is independent of ApoB.

## Discussion

We applied PCA on 168 ^1^H NMR-based metabolomic measures in 56,712 UKB participants and identified 6 main PCs representing independent metabolomic profiles, with a cumulative explained variance of 88%. We subsequently used multivariable-adjusted Cox regression and large-scale multicohort MR analyses to examine the cardiometabolic risks associated with these PCs. We found that PC1 (characterized by higher levels of ApoB and ApoB-containing lipoproteins) was associated with higher risk of CAD, and PC3 (characterized by lower levels of most ApoB-containing lipoproteins and higher levels of HDL particles) was associated with lower risk of CAD. Notably, PC4 (characterized by lower levels of small HDL particles and higher levels of very large HDL particles) was also associated with higher risk of CAD. PC5 (characterized by higher levels of amino acids) was associated with the risk of T2D and ISTR. Our findings with PC1 and PC3, which mainly captured higher and lower ApoB-associated lipoproteins, respectively, in relation to CAD are in line with previous findings that ApoB-containing lipoproteins drive atherogenic cardiovascular disease (1, 2, 43, 44). PC4 was also associated with CAD, but this association seemed independent from ApoB. Based on the loading values (Figure 2), the higher PC4-related risk of CAD is associated with a very specific HDL size distribution characterized by lower levels of small HDL particles and higher levels of large HDL particles. These analyses provide evidence for a potential (causal) association of HDL particles/composition with CAD, independent of ApoB.

It has previously been suggested that clinically measured HDL-C may not capture the protective effects of HDL on CAD (45). Hypotheses on HDL function have been proposed, suggesting that the protective properties of HDL, such as antioxidant effects, removal of cellular cholesterol and production of nitric oxide, may depend on specific HDL sub-particle characteristics and cannot reliably be estimated via the simple measurement of HDL-C (46, 47). Accordingly, in line with our observations, small HDL was found to have atheroprotective effects on macrophages and endothelial cells (47). Reduced hepatic scavenger receptor class BI (SR-BI) function was found to be associated with impaired reverse cholesterol transport (RCT), and participants with SR-BI deficiency had an increased risk of coronary heart disease despite increased HDL-C levels (48). Presumably, this increased HDL-C is caused by the accumulation of cholesterol loaded large HDL particles that cannot be cleared via SR-BI by the liver. In line with this interpretation, the loadings of PC4 indicated higher levels of large HDL particles associated with increased CAD risk. These and our data thus provide evidence for the hypothesis that for HDL-targeted therapy to be effective in prevention of CAD, higher levels of small HDL particles and lower levels of large HDL particles are warranted. Treatment with CETP inhibitors results in the opposite effect on the HDL profile (49-51), which may therefore at least partly explain the clinical failure of these drugs thus far.

In addition, our study found that genetically-influenced PC3 (characterized by lower levels of most ApoB-containing lipoproteins and higher levels of HDL particles) was also associated with a lower risk of ISTR. Although not all studies support an increased risk of ISTR by elevated LDL-C (52, 53), LDL-C levels were found to increase the risk of the large artery stroke subtype in recent MR studies (54, 55). Moreover, statins remain one of the main strategies to prevent ISTR (56). ApoB has also been suggested as the predominant risk factor for the risk of ISTR (57). In addition, genetically predicted HDL-C was also found to possibly decrease the risk of ISTR, particularly small vessel stroke (55, 58). Nevertheless, although no association with ISTR was demonstrated for PC1, our study provides evidence for a role of dyslipidaemia in the development of ISTR.

While cardiovascular diseases and T2D share some underlying risk factors, their relationships with lipoprotein metabolism was found to be different (59). A lipoprotein profile, consisting of higher levels of large VLDL particles and small LDL particles, lower levels of large HDL particles, smaller LDL and HDL particle size, and larger VLDL particle size, was found to be associated with incident diabetes (60-63). In the present study, the opposite of this risk lipoprotein profile was seen in PC2. Therefore, as expected, our study found that PC2 was associated with lower risk of T2D but seemingly with a weak effect in the MR analyses.

We found that PC5, mainly characterised by higher levels of amino acids, is a risk factor for T2D and ISTR. Multiple prospective analyses have observed associations of specific amino acids with increased risk (branched chain amino acids [BCAAs], alanine, phenylalanine) or decreased risk (glutamine and glycine) of T2D (64-67). The metabolism of amino acids has been shown to be crucial for the development and progression of ISTR (68). For example, higher levels of BCAAs and glutamate have consistently been reported to contribute to an increased risk of ISTR (69, 70).

There are three main strengths of the present study. First, the large number of ^1^H-NMR-based metabolomic measures in a very large number of disease free individuals, especially various lipids and lipoprotein fractions, enabled thorough description of the interrelationship among metabolomic measures and the identification of specific profiles. Second, for MR analyses, a large-scale multicohort design was used, which provided ample power. Third, the two-sample MR study design reduced bias by non-overlapping samples between gene-exposure association analyses and gene-outcome association analyses. In addition, pleotropic effects were avoided by excluding overlapping SNPs among PCs.

There are also several limitations to be considered. The metabolomic measures from UKB are from non-fasting samples. Non-fasting may result in measurements of lipids and lipoproteins that are not representative for average daily levels, especially TG and LDL-C (71). However, recent studies suggested that fasting is not routinely required for risk analysis of lipid profiles, and that the measurement of ApoB is stable with or without fasting (71-74). Additionally, the exposures were assessed based on PCs that represented standardised composite traits, so the estimates cannot be interpreted as effects of specific metabolomic biomarkers per unit change, which may decrease the direct clinical value and requires further research.

In conclusion, the present study, based on independent profiles of metabolomic measures, not only confirmed the effect of ApoB-containing lipoproteins on CAD, but also revealed the existence of an alternative ApoB-independent metabolomic profile associated with CAD risk, providing evidence for the potential role of HDL in the development of CAD. Furthermore, our findings support the notion that lipids, lipoproteins and amino acids are important risk factors for the development of T2D and ISTR. More research focusing on specific metabolomic measures is needed to investigate their specific causal role in the development of cardiometabolic disease.

## Data Availability

Data used in the multivariable-adjusted analyses will be made available upon request in adherence with transparency conventions in medical research and through reasonable requests to the corresponding author. Data used in the MR analyses are all publicly available provided in the article or via the corresponding consortium.

## Acknowledgements

The present study has been conducted using the UK Biobank Resource (Application Number 56340) that is available to researchers. The authors acknowledge the participants and investigators of all consortia that contributed summary statistics data, including the UK Biobank, CARDIoGRAMplusC4D, HERMES consortium, and the FinnGen study.

## References

1. Ference BA, Ginsberg HN, Graham I, Ray KK, Packard CJ, Bruckert E, et al. Low-density lipoproteins cause atherosclerotic cardiovascular disease. 1. Evidence from genetic, epidemiologic, and clinical studies. A consensus statement from the European Atherosclerosis Society Consensus Panel. Eur Heart J. 2017;38(32):2459–72.

2. Ibi D, Blauw LL, Noordam R, Dolle MET, Jukema JW, Rosendaal FR, et al. Triglyceride-lowering LPL alleles combined with LDL-C-lowering alleles are associated with an additively improved lipoprotein profile. Atherosclerosis. 2021;328:144–52.

3. Lotta LA, Stewart ID, Sharp SJ, Day FR, Burgess S, Luan J, et al. Association of Genetically Enhanced Lipoprotein Lipase-Mediated Lipolysis and Low-Density Lipoprotein Cholesterol-Lowering Alleles With Risk of Coronary Disease and Type 2 Diabetes. JAMA Cardiol. 2018;3(10):957–66.

4. Sniderman AD, Thanassoulis G, Glavinovic T, Navar AM, Pencina M, Catapano A, et al. Apolipoprotein B Particles and Cardiovascular Disease: A Narrative Review. JAMA Cardiol. 2019;4(12):1287–95.

5. Richardson TG, Sanderson E, Palmer TM, Ala-Korpela M, Ference BA, Davey Smith G, et al. Evaluating the relationship between circulating lipoprotein lipids and apolipoproteins with risk of coronary heart disease: A multivariable Mendelian randomisation analysis. PLoS Med. 2020;17(3):e1003062.

6. Ference BA, Kastelein JJP, Ray KK, Ginsberg HN, Chapman MJ, Packard CJ, et al. Association of Triglyceride-Lowering LPL Variants and LDL-C-Lowering LDLR Variants With Risk of Coronary Heart Disease. JAMA. 2019;321(4):364–73.

7. Marston NA, Giugliano RP, Melloni GEM, Park JG, Morrill V, Blazing MA, et al. Association of Apolipoprotein B-Containing Lipoproteins and Risk of Myocardial Infarction in Individuals With and Without Atherosclerosis: Distinguishing Between Particle Concentration, Type, and Content. JAMA Cardiol. 2022;7(3):250–6.

8. The Emerging Risk Factors Collaboration. Major Lipids, Apolipoproteins, and Risk of Vascular Disease. JAMA. 2009;302(18):1993–2000.

9. Gofman JW, Young W, Tandy R. Ischemic Heart Disease, Atherosclerosis, and Longevity. Circulation. 1966;34(4):679–97.

10. Voight BF, Peloso GM, Orho-Melander M, Frikke-Schmidt R, Barbalic M, Jensen MK, et al. Plasma HDL cholesterol and risk of myocardial infarction: a mendelian randomisation study. The Lancet. 2012;380(9841):572–80.

11. Frikke-Schmidt R, Nordestgaard BG, Stene MCA, Sethi AA, Remaley AT, Schnohr P, et al. Association of Loss-of-Function Mutations in the ABCA1 Gene With High-Density Lipoprotein Cholesterol Levels and Risk of Ischemic Heart Disease. JAMA. 2008;299(21):2524–32.

12. Schwartz GG, Olsson AG, Abt M, Ballantyne CM, Barter PJ, Brumm J, et al. Effects of dalcetrapib in patients with a recent acute coronary syndrome. N Engl J Med. 2012;367(22):2089–99.

13. Lincoff AM, Nicholls SJ, Riesmeyer JS, Barter PJ, Brewer HB, Fox KAA, et al. Evacetrapib and Cardiovascular Outcomes in High-Risk Vascular Disease. N Engl J Med. 2017;376(20):1933–42.

14. Soininen P, Kangas AJ, Wurtz P, Suna T, Ala-Korpela M. Quantitative serum nuclear magnetic resonance metabolomics in cardiovascular epidemiology and genetics. Circ Cardiovasc Genet. 2015;8(1):192–206.

15. Iida M, Harada S, Takebayashi T. Application of Metabolomics to Epidemiological Studies of Atherosclerosis and Cardiovascular Disease. J Atheroscler Thromb. 2019;26(9):747–57.

16. Cavus E, Karakas M, Ojeda FM, Kontto J, Veronesi G, Ferrario MM, et al. Association of Circulating Metabolites With Risk of Coronary Heart Disease in a European Population: Results From the Biomarkers for Cardiovascular Risk Assessment in Europe (BiomarCaRE) Consortium. JAMA Cardiol. 2019;4(12):1270–9.

17. Wurtz P, Kangas AJ, Soininen P, Lehtimaki T, Kahonen M, Viikari JS, et al. Lipoprotein subclass profiling reveals pleiotropy in the genetic variants of lipid risk factors for coronary heart disease: a note on Mendelian randomization studies. J Am Coll Cardiol. 2013;62(20):1906–8.

18. Sudlow C, Gallacher J, Allen N, Beral V, Burton P, Danesh J, et al. UK biobank: an open access resource for identifying the causes of a wide range of complex diseases of middle and old age. PLoS Med. 2015;12(3):e1001779.

19. Lawlor DA, Tilling K, Davey Smith G. Triangulation in aetiological epidemiology. Int J Epidemiol. 2016;45(6):1866–86.

20. Bycroft C, Freeman C, Petkova D, Band G, Elliott LT, Sharp K, et al. The UK Biobank resource with deep phenotyping and genomic data. Nature. 2018;562(7726):203–9.

21. Wurtz P, Kangas AJ, Soininen P, Lawlor DA, Davey Smith G, Ala-Korpela M. Quantitative Serum Nuclear Magnetic Resonance Metabolomics in Large-Scale Epidemiology: A Primer on -Omic Technologies. Am J Epidemiol. 2017;186(9):1084–96.

22. Jolliffe IT, Cadima J. Principal component analysis: a review and recent developments. Philos Trans A Math Phys Eng Sci. 2016;374(2065):20150202.

23. Smith GD, Ebrahim S. ‘Mendelian randomization’: can genetic epidemiology contribute to understanding environmental determinants of disease? Int J Epidemiol. 2003;32(1):1–22.

24. Davies NM, Holmes MV, Davey Smith G. Reading Mendelian randomisation studies: a guide, glossary, and checklist for clinicians. BMJ. 2018;362:k601.

25. Pierce BL, Burgess S. Efficient design for Mendelian randomization studies: subsample and 2-sample instrumental variable estimators. Am J Epidemiol. 2013;178(7):1177–84.

26. Burgess S, Scott RA, Timpson NJ, Davey Smith G, Thompson SG, Consortium E-I. Using published data in Mendelian randomization: a blueprint for efficient identification of causal risk factors. Eur J Epidemiol. 2015;30(7):543–52.

27. Burgess S, Butterworth A, Thompson SG. Mendelian randomization analysis with multiple genetic variants using summarized data. Genet Epidemiol. 2013;37(7):658–65.

28. UK10K Consortium, Walter K, Min JL, Huang J, Crooks L, Memari Y, et al. The UK10K project identifies rare variants in health and disease. Nature. 2015;526(7571):82–90.

29. 1000 Genomes Project Consortium, Auton A, Brooks LD, Durbin RM, Garrison EP, Kang HM, et al. A global reference for human genetic variation. Nature. 2015;526(7571):68–74.

30. McCarthy S, Das S, Kretzschmar W, Delaneau O, Wood AR, Teumer A, et al. A reference panel of 64,976 haplotypes for genotype imputation. Nat Genet. 2016;48(10):1279–83.

31. Westerman KE, Pham DT, Hong L, Chen Y, Sevilla-González M, Sung YJ, et al. GEM: scalable and flexible gene–environment interaction analysis in millions of samples. Bioinformatics. 2021;37(20):3514–20.

32. Hemani G, Zheng J, Elsworth B, Wade K, Haberland V, Baird D, et al. The MR-Base platform supports systematic causal inference across the human phenome. eLife. 2018;7:e34408.

33. Shim H, Chasman DI, Smith JD, Mora S, Ridker PM, Nickerson DA, et al. A multivariate genome-wide association analysis of 10 LDL subfractions, and their response to statin treatment, in 1868 Caucasians. PLoS One. 2015;10(4):e0120758.

34. Nikpay M, Goel A, Won HH, Hall LM, Willenborg C, Kanoni S, et al. A comprehensive 1,000 Genomes-based genome-wide association meta-analysis of coronary artery disease. Nat Genet. 2015;47(10):1121–30.

35. Mahajan A, Taliun D, Thurner M, Robertson NR, Torres JM, Rayner NW, et al. Fine-mapping type 2 diabetes loci to single-variant resolution using high-density imputation and islet-specific epigenome maps. Nat Genet. 2018;50(11):1505–13.

36. Malik R, Chauhan G, Traylor M, Sargurupremraj M, Okada Y, Mishra A, et al. Multiancestry genome-wide association study of 520,000 subjects identifies 32 loci associated with stroke and stroke subtypes. Nat Genet. 2018;50(4):524–37.

37. Kurki MI, Karjalainen J, Palta P, Sipilä TP, Kristiansson K, Donner K, et al. FinnGen: Unique genetic insights from combining isolated population and national health register data. medRxiv. 2022:2022.03.03.22271360.

38. Hoaglin DC. We know less than we should about methods of meta-analysis. Res Synth Methods. 2015;6(3):287–9.

39. Bowden J, Davey Smith G, Haycock PC, Burgess S. Consistent Estimation in Mendelian Randomization with Some Invalid Instruments Using a Weighted Median Estimator. Genet Epidemiol. 2016;40(4):304–14.

40. Burgess S, Thompson SG. Interpreting findings from Mendelian randomization using the MR-Egger method. Eur J Epidemiol. 2017;32(5):377–89.

41. Bowden J, Davey Smith G, Burgess S. Mendelian randomization with invalid instruments: effect estimation and bias detection through Egger regression. Int J Epidemiol. 2015;44(2):512–25.

42. Kaiser HF. The Application of Electronic Computers to Factor Analysis. Educational and Psychological Measurement. 1960;20(1):141–51.

43. Visseren FLJ, Mach F, Smulders YM, Carballo D, Koskinas KC, Back M, et al. 2021 ESC Guidelines on cardiovascular disease prevention in clinical practice. Eur Heart J. 2021;42(34):3227–337.

44. Mach F, Baigent C, Catapano AL, Koskinas KC, Casula M, Badimon L, et al. 2019 ESC/EAS Guidelines for the management of dyslipidaemias: lipid modification to reduce cardiovascular risk. Eur Heart J. 2020;41(1):111–88.

45. McGarrah RW, Craig DM, Haynes C, Dowdy ZE, Shah SH, Kraus WE. High-density lipoprotein subclass measurements improve mortality risk prediction, discrimination and reclassification in a cardiac catheterization cohort. Atherosclerosis. 2016;246:229–35.

46. Rader DJ, Hovingh GK. HDL and cardiovascular disease. The Lancet. 2014;384(9943):618–25.

47. Calabresi L, Gomaraschi M, Simonelli S, Bernini F, Franceschini G. HDL and atherosclerosis: Insights from inherited HDL disorders. Biochim Biophys Acta. 2015;1851(1):13–8.

48. Zanoni P, Khetarpal SA, Larach DB, Hancock-Cerutti WF, Millar JS, Cuchel M, et al. Rare variant in scavenger receptor BI raises HDL cholesterol and increases risk of coronary heart disease. Science. 2016;351(6278):1166–71.

49. Samedy L-A, Ryan GJ, Superko RH, Momary KM. CETP genotype and concentrations of HDL and lipoprotein subclasses in African–American men. Future Cardiology. 2019;15(3):187–95.

50. Nicholls SJ, Brewer HB, Kastelein JJP, Krueger KA, Wang M-D, Shao M, et al. Effects of the CETP Inhibitor Evacetrapib Administered as Monotherapy or in Combination With Statins on HDL and LDL Cholesterol: A Randomized Controlled Trial. JAMA. 2011;306(19):2099–109.

51. Kastelein JJP, van Leuven SI, Burgess L, Evans GW, Kuivenhoven JA, Barter PJ, et al. Effect of Torcetrapib on Carotid Atherosclerosis in Familial Hypercholesterolemia. New England Journal of Medicine. 2007;356(16):1620–30.

52. Yaghi S, Elkind MS. Lipids and Cerebrovascular Disease: Research and Practice. Stroke. 2015;46(11):3322–8.

53. Valdes-Marquez E, Parish S, Clarke R, Stari T, Worrall BB, ISGC MCot, et al. Relative effects of LDL-C on ischemic stroke and coronary disease: A Mendelian randomization study. Neurology. 2019;92(11):e1176–e87.

54. Hindy G, Engstrom G, Larsson SC, Traylor M, Markus HS, Melander O, et al. Role of Blood Lipids in the Development of Ischemic Stroke and its Subtypes: A Mendelian Randomization Study. Stroke. 2018;49(4):820–7.

55. Georgakis MK, Gill D. Mendelian Randomization Studies in Stroke: Exploration of Risk Factors and Drug Targets With Human Genetic Data. Stroke. 2021;52(9):2992–3003.

56. Kernan WN, Ovbiagele B, Black HR, Bravata DM, Chimowitz MI, Ezekowitz MD, et al. Guidelines for the prevention of stroke in patients with stroke and transient ischemic attack: a guideline for healthcare professionals from the American Heart Association/American Stroke Association. Stroke. 2014;45(7):2160–236.

57. Yuan S, Tang B, Zheng J, Larsson SC. Circulating Lipoprotein Lipids, Apolipoproteins and Ischemic Stroke. Ann Neurol. 2020;88(6):1229–36.

58. Georgakis MK, Malik R, Anderson CD, Parhofer KG, Hopewell JC, Dichgans M. Genetic determinants of blood lipids and cerebral small vessel disease: role of high-density lipoprotein cholesterol. Brain. 2020;143(2):597–610.

59. Goodarzi MO, Rotter JI. Genetics Insights in the Relationship Between Type 2 Diabetes and Coronary Heart Disease. Circ Res. 2020;126(11):1526–48.

60. Hodge AM, Jenkins AJ, English DR, O’Dea K, Giles GG. NMR-determined lipoprotein subclass profile predicts type 2 diabetes. Diabetes Res Clin Pract. 2009;83(1):132–9.

61. Mora S, Otvos JD, Rosenson RS, Pradhan A, Buring JE, Ridker PM. Lipoprotein particle size and concentration by nuclear magnetic resonance and incident type 2 diabetes in women. Diabetes. 2010;59(5):1153–60.

62. Mackey RH, Mora S, Bertoni AG, Wassel CL, Carnethon MR, Sibley CT, et al. Lipoprotein particles and incident type 2 diabetes in the multi-ethnic study of atherosclerosis. Diabetes Care. 2015;38(4):628–36.

63. Shalaurova I, Connelly MA, Garvey WT, Otvos JD. Lipoprotein insulin resistance index: a lipoprotein particle-derived measure of insulin resistance. Metab Syndr Relat Disord. 2014;12(8):422–9.

64. Wang TJ, Larson MG, Vasan RS, Cheng S, Rhee EP, McCabe E, et al. Metabolite profiles and the risk of developing diabetes. Nat Med. 2011;17(4):448–53.

65. Floegel A, Stefan N, Yu Z, Muhlenbruch K, Drogan D, Joost HG, et al. Identification of serum metabolites associated with risk of type 2 diabetes using a targeted metabolomic approach. Diabetes. 2013;62(2):639–48.

66. Tillin T, Hughes AD, Wang Q, Wurtz P, Ala-Korpela M, Sattar N, et al. Diabetes risk and amino acid profiles: cross-sectional and prospective analyses of ethnicity, amino acids and diabetes in a South Asian and European cohort from the SABRE (Southall And Brent REvisited) Study. Diabetologia. 2015;58(5):968–79.

67. Morze J, Wittenbecher C, Schwingshackl L, Danielewicz A, Rynkiewicz A, Hu FB, et al. Metabolomics and Type 2 Diabetes Risk: An Updated Systematic Review and Meta-analysis of Prospective Cohort Studies. Diabetes Care. 2022;45(4):1013–24.

68. Ke C, Pan CW, Zhang Y, Zhu X, Zhang Y. Metabolomics facilitates the discovery of metabolic biomarkers and pathways for ischemic stroke: a systematic review. Metabolomics. 2019;15(12):152.

69. Ruiz-Canela M, Toledo E, Clish CB, Hruby A, Liang L, Salas-Salvado J, et al. Plasma Branched-Chain Amino Acids and Incident Cardiovascular Disease in the PREDIMED Trial. Clin Chem. 2016;62(4):582–92.

70. Holmes MV, Millwood IY, Kartsonaki C, Hill MR, Bennett DA, Boxall R, et al. Lipids, Lipoproteins, and Metabolites and Risk of Myocardial Infarction and Stroke. J Am Coll Cardiol. 2018;71(6):620–32.

71. Cartier LJ, Collins C, Lagace M, Douville P. Comparison of fasting and non-fasting lipid profiles in a large cohort of patients presenting at a community hospital. Clin Biochem. 2018;52:61–6.

72. Mora S, Chang CL, Moorthy MV, Sever PS. Association of Nonfasting vs Fasting Lipid Levels With Risk of Major Coronary Events in the Anglo-Scandinavian Cardiac Outcomes Trial-Lipid Lowering Arm. JAMA Intern Med. 2019;179(7):898–905.

73. Nordestgaard BG, Langsted A, Mora S, Kolovou G, Baum H, Bruckert E, et al. Fasting is not routinely required for determination of a lipid profile: clinical and laboratory implications including flagging at desirable concentration cut-points-a joint consensus statement from the European Atherosclerosis Society and European Federation of Clinical Chemistry and Laboratory Medicine. Eur Heart J. 2016;37(25):1944–58.

74. Nordestgaard BG. A Test in Context: Lipid Profile, Fasting Versus Nonfasting. Journal of the American College of Cardiology. 2017;70(13):1637–46.

