## Supplemental Figures for "Causal evidence for an ApoB-independent metabolomic risk profile associated with coronary artery disease"

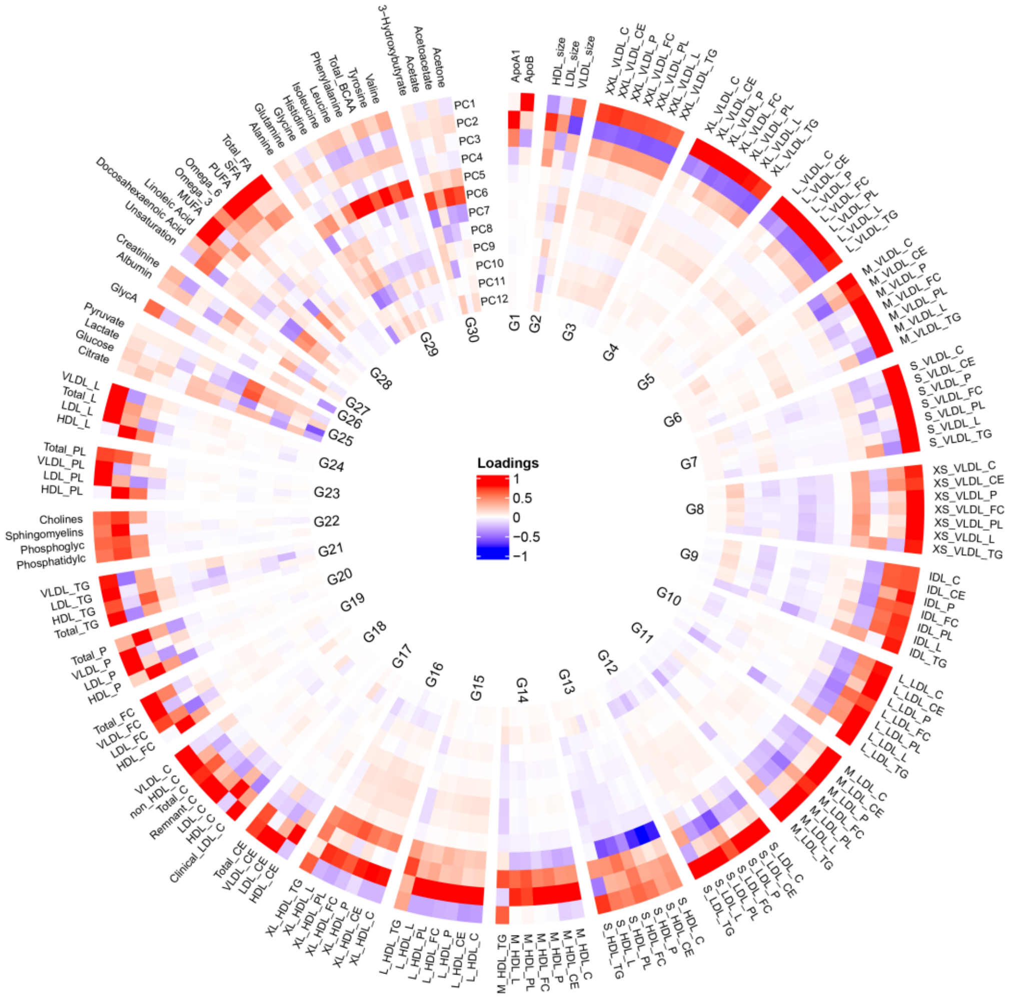


**Groups:**

| | | | | |
|---|---|---|---|---|
| G1 Apolipoproteins | G7 Small VLDL | G13 Small HDL | G19 Free cholesterol | G25 Glycolysis related metabolites |
| G2 Lipoprotein particle sizes | G8 Very small VLDL | G14 Medium HDL | G20 Lipoprotein particle concentrations | G26 Inflammation |
| G3 Extremely large VLDL | G9 IDL | G15 Large HDL | G21 Triglycerides | G27 Fluid balance |
| G4 Very large VLDL | G10 Large LDL | G16 Very large HDL | G22 Other lipids | G28 Fatty acids |
| G5 Large VLDL | G11 Medium LDL | G17 Cholesteryl esters | G23 Phospholipids | G29 Amino acids |
| G6 Medium VLDL | G12 Small LDL | G18 Cholesterol | G24 Total lipids | G30 Ketone bodies |

**Figure S1. A circular loading plot of the first twelve principal components.** From the outside to the inside, each circle represents the correlations between the respective principle components (PC) and 168 metabolomic measures. All metabolomic measures were divided into 30 groups and shown in clockwise order. Red or blue colour indicates the increase or decrease of metabolomic measures in PCs.

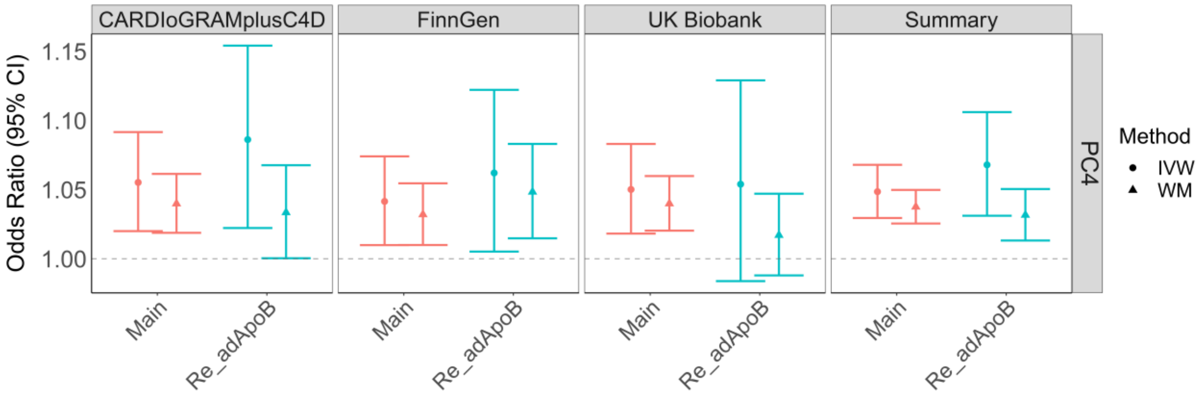


**Figure S2. Odds ratios (95% CI) for the risk of CAD/CHD per one-SD increase in PC4 and residual-PC4 by inverse-variance weighted method and weighted median method.** Four column panel showed the ORs[95% CI] from three separate data sources, and the pooled ORs labelled 'Summary' from the fixed effects meta-analysis. The estimated associations between CAD and PC4 in the main analysis were marked on the X-axis with "Main" and in red, and the estimated associations between CAD and residual-PC4 after adjustment for ApoB were marked on the X-axis with "Re_adApoB" and in blue.
